# Low Positive Social Determinants of Health Exposure Amplifies the Association between Allostatic Load and Risk of Cardiovascular Mortality in Cancer and Non-Cancer Patients

**DOI:** 10.1101/2023.11.22.23298933

**Authors:** Aditya Bhave, Ritu Reddy, Justin X. Moore, Priyanshu Nain, Biplab Datta, Darryl Nettles, Lakshya Seth, Stephanie Jiang, Vraj Patel, Sarah Malik, Neal Weintraub, Sadeer G Al-Kindi, Sarju Ganatra, Sourbha Dani, Javier Gomez-Valencia, Xiaoling Wang, Avirup Guha

**Affiliations:** Department of Medicine, Medical College of Georgia, Augusta, GA, United States; Center for Healthy Equity Transformation, University of Kentucky College of Medicine, Lexington, KY, United States; Department of Population Health Sciences, Medical College of Georgia, Augusta, GA, United States; Cardio-Oncology Program, Medical College of Georgia, Augusta, GA, United States; Cancer Informatics, Seidman Cancer Center at University Hospitals of Cleveland, Cleveland, OH, United States; Lahey Hospital and Medical Center, Beth Israel Laney Health, Burlington, MA, United States; Cook County Health, Chicago, IL, United States

**Keywords:** Allostatic Load, Social Determinants, Cardiovascular Disease

## Abstract

**Background:** High allostatic load (AL) is associated with an increased risk of cardiovascular death (CVD), but little is known about how social determinants moderate this relationship.

**Methods:** We conducted a retrospective cohort analysis of the National Health and Nutrition Examination Survey years 1999-2010 linked with the National Death Index. We fit age, race, and sex-adjusted Fine & Gray models to calculate sub-distribution hazard ratios (SHR) of CVD among adults exposed to high versus low levels of positive social determinants of health (PSDOH), stratified by high and low AL status.

**Results:** Among 22,775 participants, 1,939 (8.5%) had a cancer history. In the full cohort, low PSDOH was associated with 38% increased risk of CVD among high AL adults (SHR: 1.38, 95% CI: 1.22 - 1.56) and 57% increased risk among low AL adults (SHR: 1.57, 95% CI: 1.32 - 1.87). Among adults with no cancer history, low PSDOH was associated with 36% increased risk among high AL adults (SHR: 1.36, 95% CI: 1.19 – 1.55) and 59% increased risk among low AL adults (SHR: 1.59, 95% CI: 1.32 – 1.92). In the cancer cohort, low PSDOH was associated with 77% increased risk among high AL adults (SHR: 1.77, 95% CI: 1.32 – 2.37) and 73% increased risk among low AL adults (SHR: 1.73, 95% CI: 1.02 – 2.94).

**Conclusions:** Low PSDOH amplified the association between AL and CVD in all cohorts, with the highest risk increases among adults with cancer history. This suggests that socioeconomic-related distress supersedes biological changes from AL in relation to CVD.

## Introduction

Cardiovascular disease is the leading cause of death from chronic disease, accounting for 1 in 3 deaths in the US (Schultz et al., 2018). Disparities in cardiovascular disease mortality (CVD) have been observed across both racial and socioeconomic gradients. Social determinants of health (SDOH) include poverty level, food security level, home ownership, household education, access to a routine place for health care, health insurance status, and household head relationship status (Connolly et al., 2022). An inverse association has been found between CVD and adversity in each of these SDOHs (Singh et al., 2015), (Sims et al., 2020) (Chang et al., 2021), (Alcalá et al., 2015).

Socioeconomic status (SES) has proven to be a strong indicator of health outcomes, partly because of chronic stress experienced by those of lower socioeconomic statuses. Allostatic load (AL) is a measure of chronic toxic stress that uses biomarkers from different physiological systems, including neural and endocrine, to quantify stress induced biological risk. High AL is associated with poorer health outcomes (Guidi et al, 2021). Allostatic load has been shown to increase with decreasing SES in the same way as CVD mortality. Neighborhood poverty, food insecurity, lack of access to health insurance and no stable home ownership were found to have a positive association with AL (Schulz et al., 2012), (Pak, Kim, 2021) (Rollston, 2020). Allostatic load has been found to be associated with negative health outcomes and increased mortality overall (Guidi et al, 2021). High AL was associated with a 22% increased risk for all-cause mortality and a 31% increased risk for CVD mortality (Parker et al., 2022).

Studies have yet to determine the effect of adverse social determinants of health as a moderator for the relationship between AL and CVD. Furthermore, existing studies have not investigated these relationships among individuals with a history of cancer to determine if cancer history affects AL, social determinants of health, and CVD associations.

## Methods

### Study Design and Participants

We performed a retrospective cohort analysis using data collected from the National Health and Nutrition Examination Survey (NHANES), a biennial survey which utilizes a multistage probability sampling design to select a representative sample of non-institutionalized US residents (Johnson et al., 2013). Details regarding NHANES data is presented in **Supplemental Methods**. Participants were considered eligible if they were over the age of 18, were not pregnant, and had available biomarker data for AL and interview and questionnaire data for positive social determinants of health (PSDOH). Participants that were missing any of these data or missing follow up time were excluded from this study (**Figure 1**). Participants who also were missing information regarding censoring or death in the NDI linkage were also excluded as they did not have adequate information to follow up.

**Figure 1:**
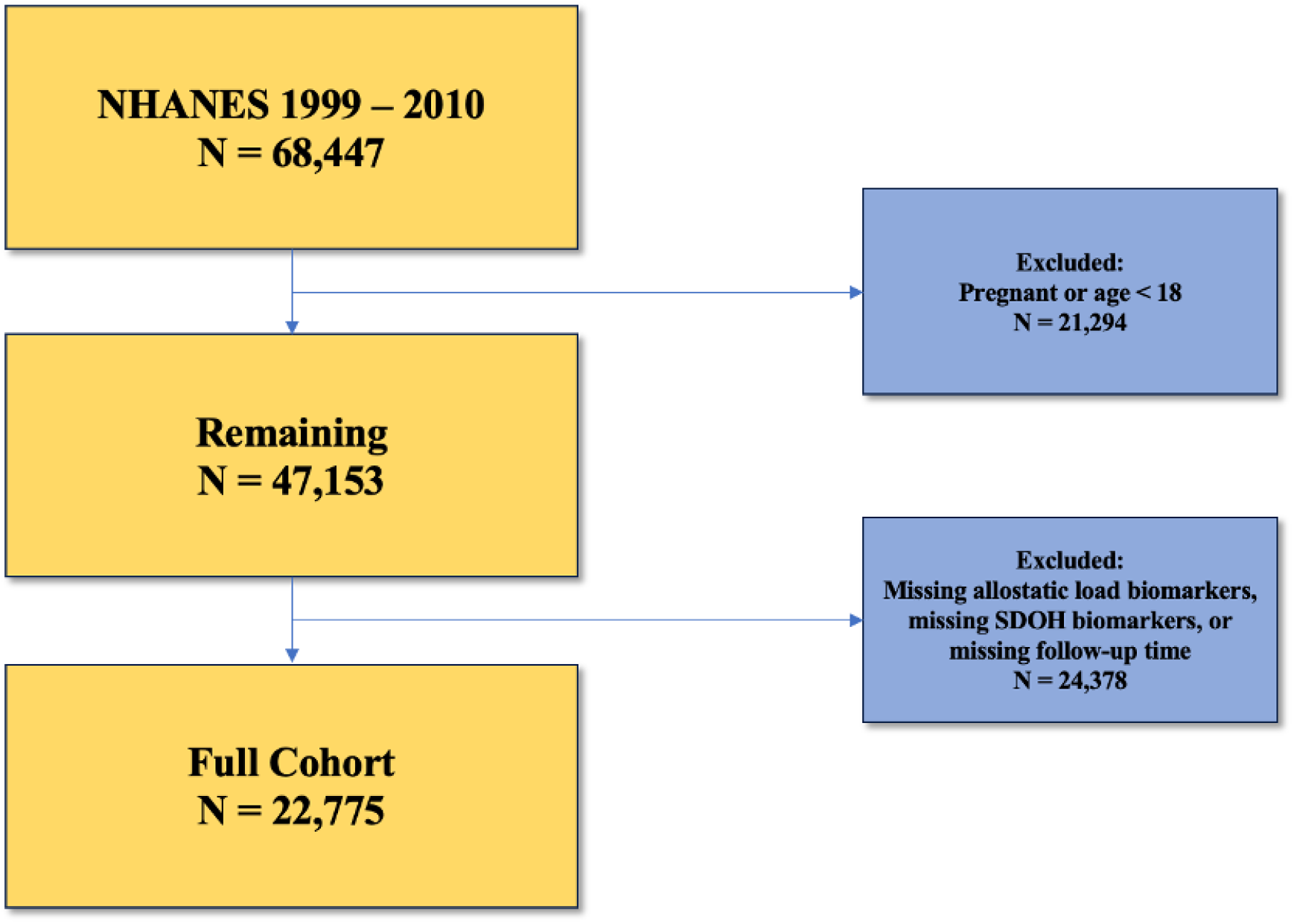
Consort Diagram: Outline of exclusion criteria applied to NHANES participants and final study population.

### Exposures of Interest: Allostatic Load and Positive Social Determinants of Health

Allostatic load (AL) has been defined in a variety of ways, but generally refers to the cumulative burden of chronic toxic stress exposure on the body and the body’s attempts to adapt (Moore et al., 2021). For this study, we defined and measured AL according to the paradigm presented by Geronimus et al. (2006) and Moore et al. (2021) that uses nine different components: body mass index (BMI), hemoglobin A1c (HbA1C), systolic blood pressure (SBP), diastolic blood pressure (DBP), serum triglycerides, serum creatinine, serum albumin, and C-reactive protein (CRP) (Geronimus et al., 2006; Moore et al., 2021). For each of these components, high-risk thresholds were determined by analyzing the gender-specific distributions of the sample. Being above the 75^th^ percentile for BMI, SBP, DBP, CRP, HbA1c, triglycerides, or creatinine or being below the 25^th^ percentile for albumin were considered high-risk (Akinyemiju et al., 2020; Frei et al., 2015). For each AL component, a score of 1 was given for each component where a participant recorded a high-risk measurement to calculate a full AL score out of a possible 9 points. Participants with a total AL score of 3 or greater were classified as having high AL, while participants with a total AL score of 2 or less were classified as having low AL (Moore et al., 2021).

Our other exposure of interest was Positive Social Determinants of Health (PSDOH). In this analysis, we utilized the framework described by Connolly et al. (2022) that matches existing data within NHANES to the WHO’s broad constructs of social determinants of health to create a novel seven-point score (Connolly et al., 2022). Economic stability was assessed by using self-reported NHANES measures of household food security, home ownership, and household poverty level index. Household Poverty level index, according to the US Health and Human Services Guidelines, is defined by the ratio of monthly family income to poverty levels, with ratios greater than 1.85 being considered favorable. Education was assessed by self-reported measure of household head educational attainment greater than a high school degree. Health care access was assessed by self-reported access to a routine place of health care (such as a doctor’s office) and by health insurance status. Social context and support was assessed by self-reported response of living with a partner. Residential environment was not assessed as data relating to the participants’ residential lives is not currently publicly available. Participants were given a score of 1 for each favorable NHANES SDOH measurement out of a total PSDOH score of 7. Participants with PSDOH scores of 4 or greater were considered as having favorable PSDOH while those with scores of 3 or less were considered as having low PSDOH. These cutoffs were chosen by examining the distribution of PSDOH scores in the sample. Being above the 75^th^ percentile in PSDOH score was considered as having high PSDOH while with scores those below this percentile cutoff were considered as having low PSDOH.

### Outcome of Interest, Cardiovascular death

The primary outcome of interest for this study was time to CVD, which was determined from the previously described linkage between NHANES and NDI. Cardiovascular-related death included both codes for cerebrovascular death as well as heart disease in the NDI linked file.

### Statistical analysis

All statistical analyses were performed using RStudio (Vienna, Austria). Descriptive statistics were generated for the three cohorts of interest – the full cohort, adults with a history of cancer, and adults without a history of cancer. This was done using the NHANES weighting guidelines described in its associated methodology handbook (CDC, 2022). Categorical variables were presented as weighted percentages and continuous variables were presented as means and associated 95% confidence intervals (CIs). Descriptive statistics were compared using the Rao-Scott-Chi-Square test for categorical variables and weighted Wald F-test for continuous variables. CVD in our study was a time-dependent variable when testing for proportional hazards assumption. Thus, we rejected proportional hazards assumptions.

To estimate relative risk of CVD between high and low PSDOH individuals stratified by high and low AL status, we fit a series of Fine & Gray models with non-CVD related mortality as the competing risk against CVD (Fine & Gray, 1999). We presented our results as subdistribution hazard ratios (SHRs) and associated 95% CIs. Our Fine & Gray models were adjusted for age, sex, and race. We also examined the multiplicative interactions of AL and PSDOH status by introducing an interaction term within our model and presented the corresponding p-values for these associations.

## Results

### Descriptive characteristics by allostatic load

**Tables 1, and supplemental Table 2** display descriptive characteristics of NHANES participants at their baseline interview stratified by AL status and cancer in the full cohort, cancer cohort (N = 1,939), and no cancer cohort (N = 20,836), respectively. Among the full cohort (N = 22,775), participants with high AL were less likely to be female (48.8% vs 51.3%, p < 0.01), and more likely to identify as Non-Hispanic Black (NHB; 12.0% vs 8.4%, p < 0.01). Participants with high AL were also more likely to have an education of less than high school (22.4% vs 16.8%, p < 0.01). Those with high AL were more likely to have had congestive heart failure (4.81% vs 1.05%, p < 0.01) and a heart attack (3.8% vs 0.8%, p < 0.01).

In the cancer cohort, participants with high AL were also more likely to identify as NHB (5.42% vs 3.49%, p < 0.01), and more likely to be in the 2^nd^ quartile of income relative to the federal poverty line (21.8% vs 16.5%, p < 0.01). They were also more likely to have had congestive heart failure and a heart attack similarly to the full cohort.

### Primary Outcome (CVD)

**Tables 2, 3 and Supplemental 6** display the adjusted SHRs and 95% confidence intervals for the association between PSDOH and CVD stratified by AL status accounting for competing risks of all-cause mortality, in the full cohort, cancer cohort, and no cancer cohort respectively.

**Table 1.**
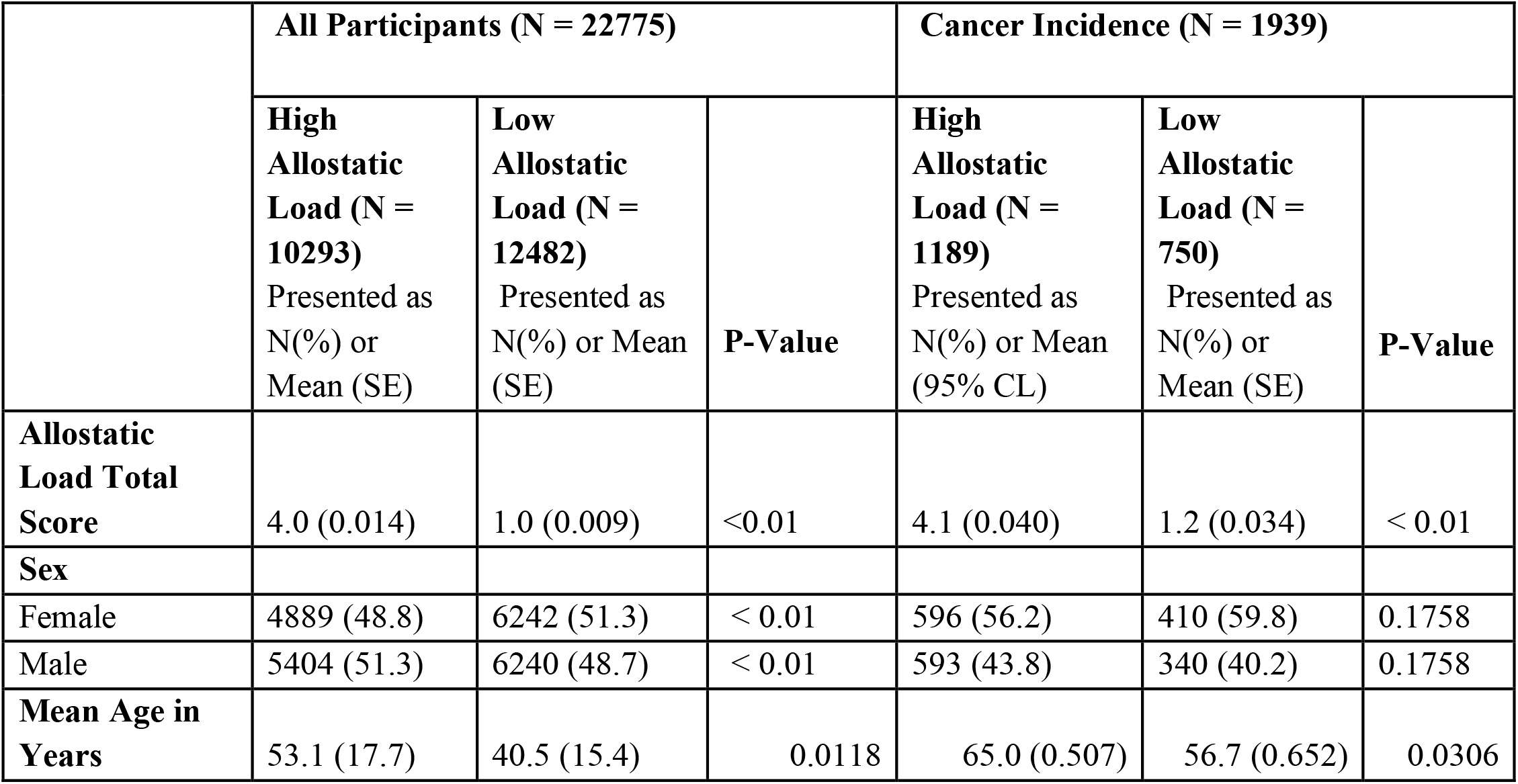
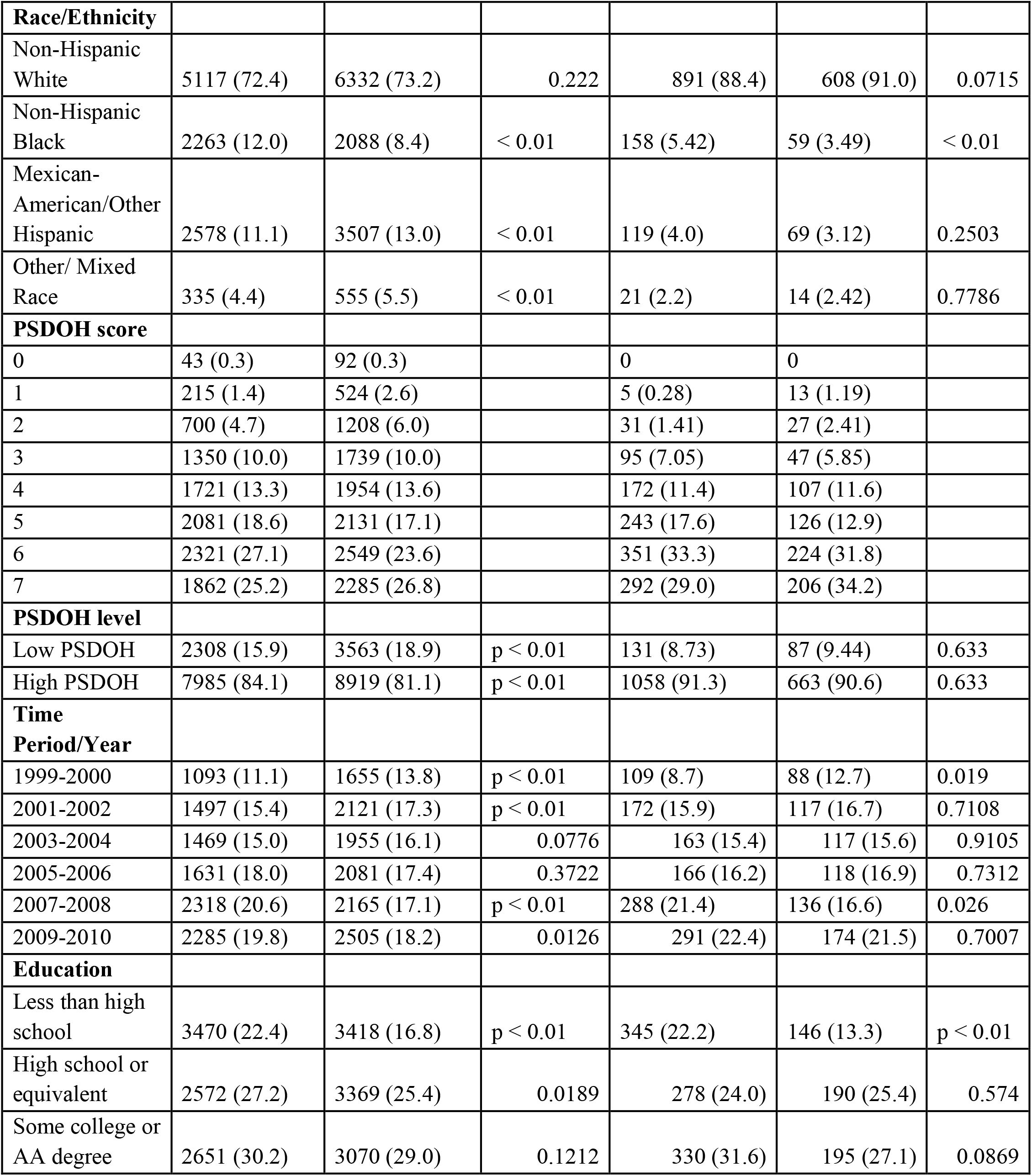
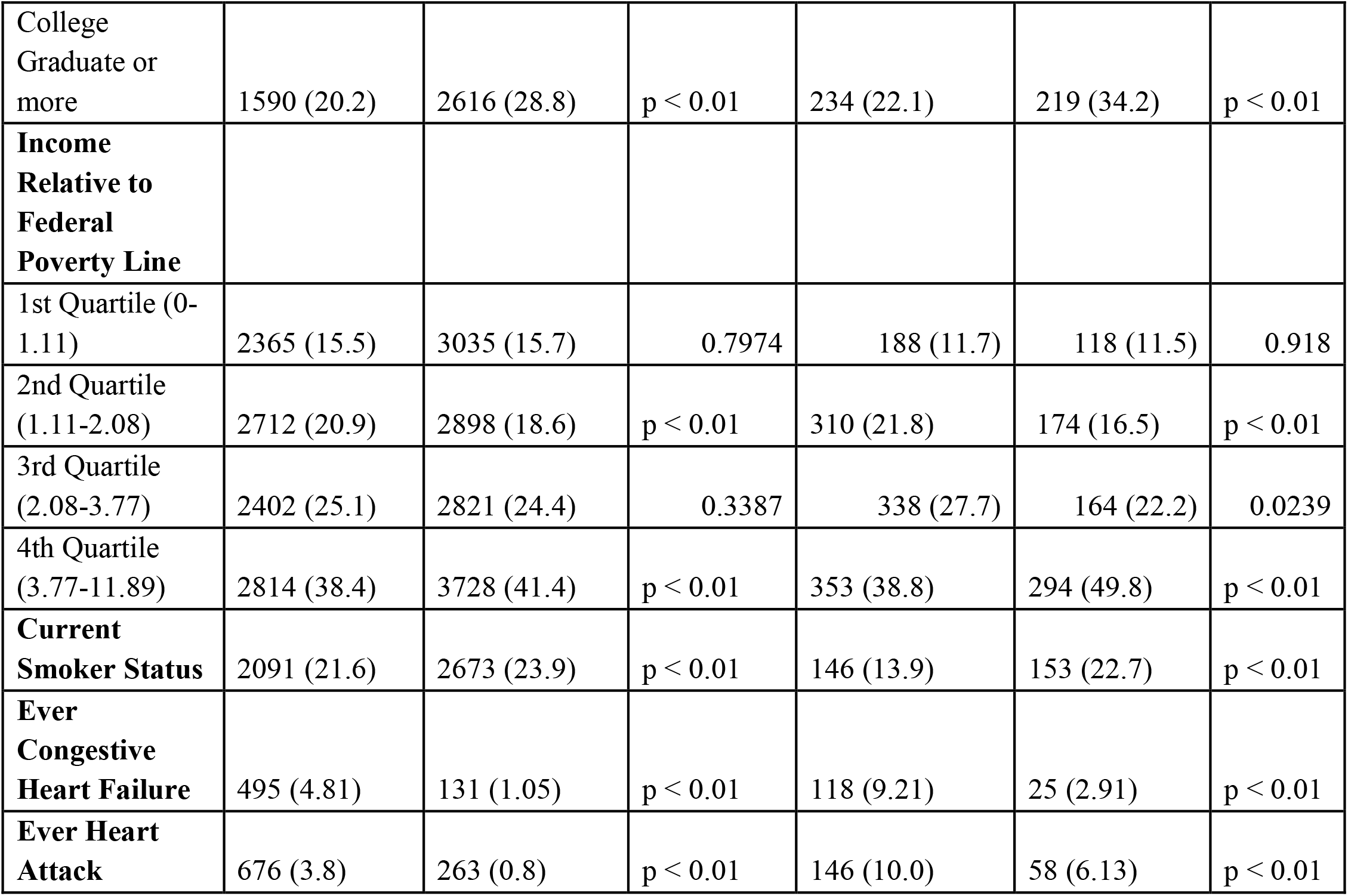
Descriptive characteristics of National Health Examination Survey (NHANES) participants stratified by AL status and cancer, years 1999-2010 with follow up through 2019.

**Table 2.**
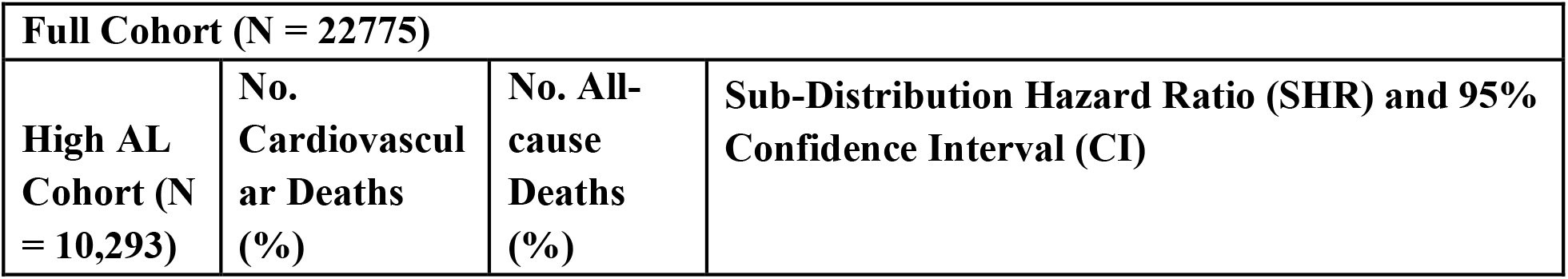
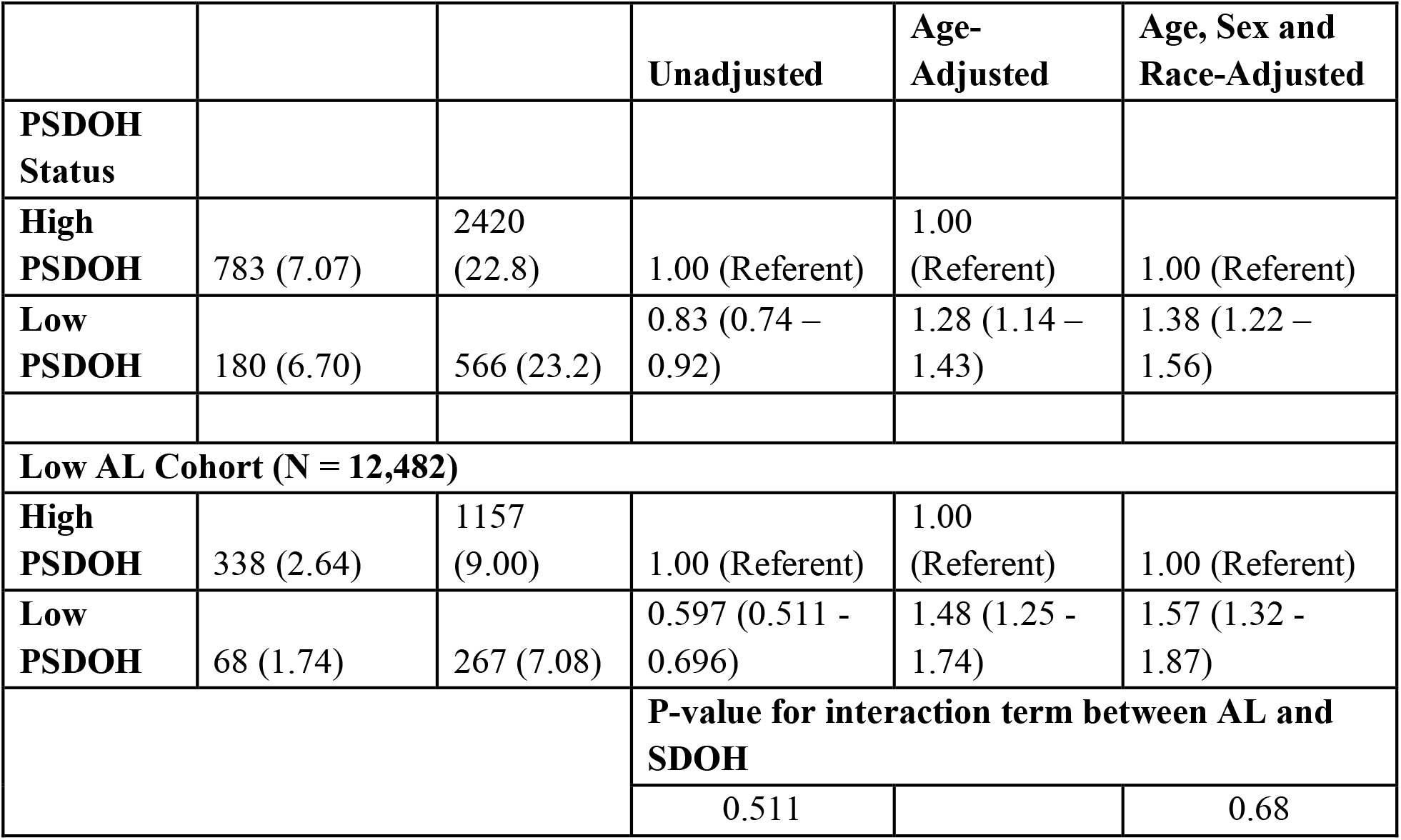
Fine & Gray method for proportional hazard models presented as Sub-Distribution Hazard ratios (SHR) and 95% confidence intervals (CI) for the association between PSDOH and CVD death stratified by AL status accounting for competing risks of all-cause mortality, among 22,775 NHANES participants with 1,369 CVD deaths, and 4,410 competing deaths.

**Table 3.**
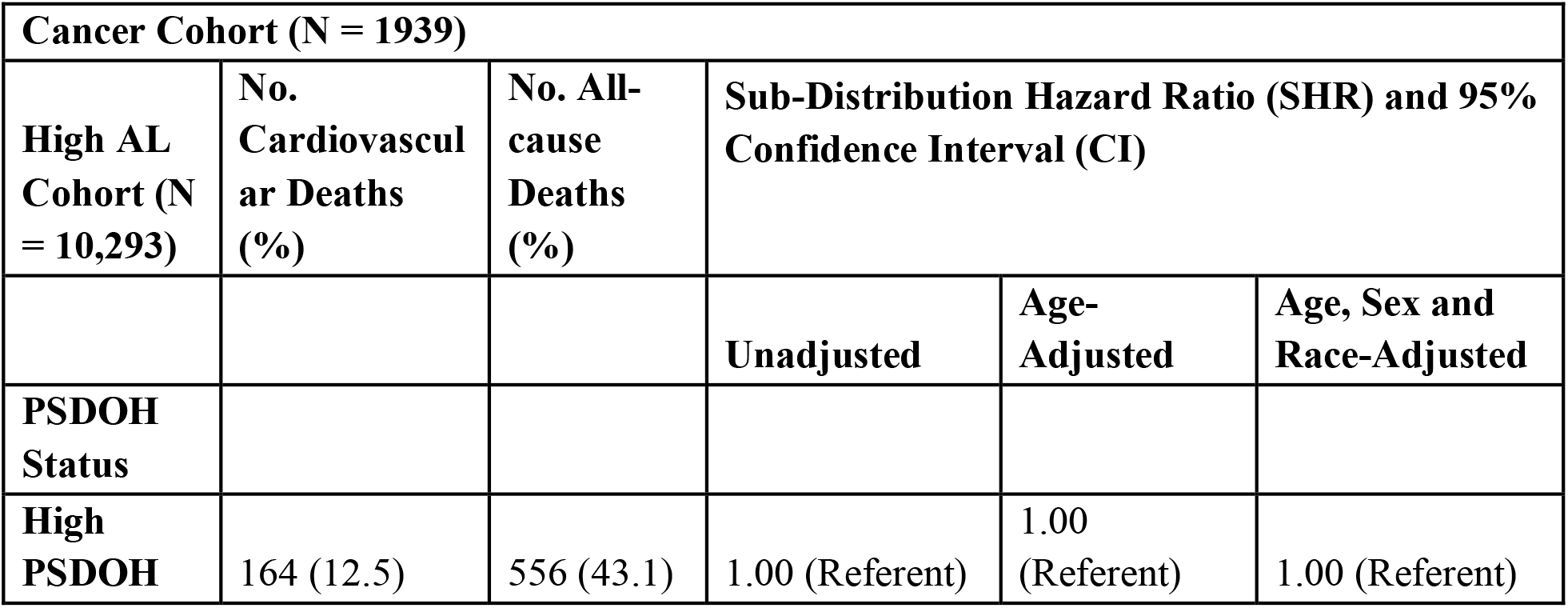
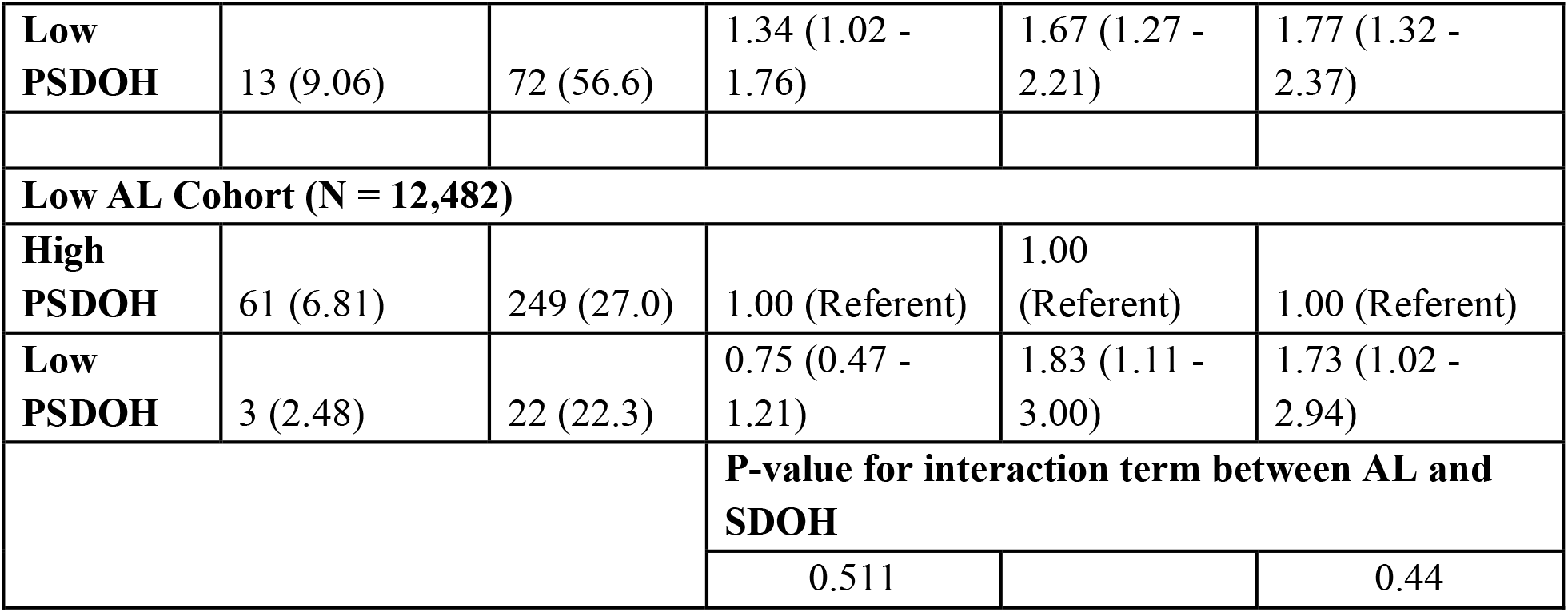
Fine & Gray method for proportional hazard models presented as Sub-Distribution Hazard ratios (SHR) and 95% confidence intervals (CI) for the association between PSDOH and CVD death among participants with a history of cancer stratified by AL status accounting for competing risks of all-cause mortality, among 1,939 NHANES participants with 241 CVD, and 899 competing deaths.

In the full cohort, cancer cohort and no cancer cohort, there were 1,369 (6.0%), 241 (12.4%) and 1,128 (5.4%) deaths from CVD. Among participants with high AL in the full cohort, low PSDOH was associated with a 38% increased risk in fully adjusted models (age, sex, and race-adjusted models, CL: 1.22-1.56). For adults with low AL, low PSDOH was associated with a 57% increased risk in fully adjusted models (age, sex, and race-adjusted models, CL: 1.32-1.87, Table 2).

Among participants with a history of cancer and high AL, low PSDOH was associated with a 34% increased risk of CVD mortality in unadjusted models (CL: 1.02-1.76) and a 77% increased risk in fully adjusted models (age, sex, and race-adjusted models, CL: 1.32-2.37). Low PSDOH in adults with a history of cancer and low AL showed a 73% increased risk for CVD mortality in fully adjusted models (CL: 1.02-2.94).

Low PSDOH in participants with no history of cancer and high AL correlated to a 36% increased risk for CVD mortality in fully adjusted models (CL: 1.19-1.55). Among adults with no history of cancer and low AL, high PSDOH was associated with a 59% increased risk of CVD mortality in fully-adjusted models (CL:1.32-1.92).

## Discussion

In our sample, considered nationally representative of non-institutionalized U.S. civilians, we found that among those with low PSDOH exposure, there was 38% increased risk of CVD among high AL adults and a 57% increased risk among low AL adults. Among adults without a history of cancer, adjusted models found that low PSDOH exposure was associated with a 36% increased risk of CVD among high AL adults and a 59% increased risk among low AL adults. Among adults with a history of cancer, adjusted models found that low PSDOH exposure was associated with a 77% increased risk of CVD among high AL adults and a 73% increased risk of CVD among low AL adults.

Our study emphasizes the great importance of social, economic, and environmental factors in the cardiovascular health of those with and without cancer. This has been corroborated by other studies examining the effects of social determinants of health on the health of NHANES sample – one such study found that among cancer survivors, an unfavorable social determinants of health profile was associated with worse cardiovascular health (Satti et al., 2023). Given the disparity in both cardiovascular health, as determined by Satti et al., and cardiovascular death outcomes determined in this study among NHANES cancer survivors, there is clearly a need for intervention at the level of clinicians, healthcare systems, and public policy to further improve cardiovascular disease outcomes among the general public.

Providers involved in the care of cardiovascular disease, especially among cancer survivors, must be educated on the links between poor social determinants of health exposure and worse health outcomes. At the clinician level, it is imperative to incorporate questions about a patient’s living conditions, educational background, insurance status, and environmental exposures during clinical assessments. This will allow for more personalized advice and interventions. For instance, dietary recommendations can be tailored to a patient’s financial capacity and local food availability and education can be provided at a level consistent with the patient’s literacy level.

On a healthcare system level, hospitals and clinics can develop partnerships with community organizations to address broader determinants like food insecurity or lack of health insurance. Programs that provide patients with resources, such farmers’ market vouchers or workshops on enrolling for public programs like Medicare and Medicaid can significantly impact the broader factors affecting their health. Healthcare systems must also strive to improve their reach and open facilities in historically disadvantaged and vulnerable areas in an effort to improve equity in health care access.

Further interventions can be taken on a policy level – increased investment in health care infrastructure by both the state and federal governments can be used to address health care access and insurance disparities. Government policy can also be utilized to address the social determinants of health themselves so that public is not affected by cardiovascular disease or cancer in the first place. Increased investment into education access, expansion of public health insurance programs, improvement of public safety, and zoning regulation to allow for more equitable access to healthy food are only some of the ways through which bodies of government can address these issues. We are just now beginning to understand the broad effects of social determinants of health on patients, disease, and health care delivery. Further research surrounding these effects as well as possible solutions should be prioritized by this nation’s universities and through bodies that provide funding for scientific research.

Given the numerous ways by which social determinants of health can affect CVD, there is a significant necessity for society-wide interventions to not only improve the care of patients afflicted with cardiovascular disease, but also address the underlying factors to prevent them from being afflicted with cardiovascular disease in the first place.

### Strengths and Limitations

This study is subject to several strengths as well as several limitations. One major advantage of the NHANES sample is its ability to be nationally representative of the U.S. civilian population. Furthermore, given the biennial nature of the NHANES survey, we were able to follow up participants for extended periods of time for a maximum of 20 years. NHANES also utilizes a standardized, tightly-controlled data collection protocol, minimizing examiner biases and reporting errors. This is particularly advantageous when studying AL in many participants over a longer time period, as it can change throughout people’s life as they age, are exposed to new stressors, and experience new life events. Prior work has demonstrated a positive correlation between AL and age (Moore et al., 2021). Increased age has also been associated with an increased risk of cardiovascular disease (Rodgers et al., 2019).

The primary limitation that we faced was having available NHANES participants with complete AL biomarker and SDOH data available to comprise our cancer cohort, which was much smaller than our no-cancer history cohort. Although the SHRs and p-value for AL and SDOH interaction revealed that we had relatively robust results, the confidence intervals generated for this group were noticeably larger than the analysis done with the no-cancer history cohort. Future studies should focus on other large, nationally representative cohorts that may have more data available on participants with a history of cancer.

The use of NHANES data itself is also subject to several limitations. The first is that data in this survey, including that of confirming a history of cancer, is self-reported. This makes the data we analyzed vulnerable to recall bias as well as underreporting or misclassification as we were not able to compare this data against physician data or medical records. Another limitation that we faced was that the NHANES data collection is cross-sectional in nature, so answers to data collection questions were provided at a single time point, while the NDI was used for longitudinal follow-up. Many of the variables in this study, such as AL, PSDOH, and even a cancer diagnosis, are subject to change in participants over time, and future studies should focus on tracking changes in AL, PSDOH, and cancer status in a large cohort. Examining these changes in individual participants in a large, longitudinal cohort can provide more robust insight into the relationship between AL, PSDOH, and cardiovascular outcomes among adults with and without cancer.

We found that once adjusting for age, race, and sex, the relationship between AL and risk of CVD was reduced, suggesting that these factors could affect both the exposures of interest (AL and PSDOH) as well as the outcome of interest (CVD).

Another limitation of our study was that proportional hazards assumptions were rejected in this study, which indicates a number of implications about the generalizability and interpretation of our results. Due to the violation of these assumptions, it is possible that the effect of our covariates and the hazard of CVD may change over time. We accounted for this by assessing the interaction between CVD and time for each of our cohorts, finding that they were not significant in each case. While this approach could not account for the time-varying effects of our covariates, we were able to understand whether the time variance of covariates represented a significant threat to the validity of our results.

## Conclusion

This study found that among participants with both high and low AL, having low positive social determinants of health exposure increased their risk for death from cardiovascular disease related causes. This relationship remained true for participants with and without a history of cancer. Future studies can find causational relationships between specific social demographics included in PSDOH and specific biological factors leading to death from CVD. Our findings further exemplify the need for healthcare that involves understanding and attempting to mitigate disparities in health and treatment due to socioeconomic factors. Studies can focus on specific approaches to lessening socioeconomic differences and how these changes affect individuals’ chronic stress and overall mortality.

## Data Availability

The NHANES database is publicly accessible (https://wwwn.cdc.gov/nchs/nhanes/Default.aspx) and contains deidentified data for all participants used in this study.

## Abbreviations

AL: allostatic load
NHANES: National Health and Nutrition Examination Survey
SHR: sub-distribution hazard ratio
CVD: Cardiovascular disease
PSDOH: positive social determinants of health
SES: socioeconomic status
BMI: body mass index
SBP: systolic blood pressure
DBP: diastolic blood pressure
HBA1C: hemoglobin
A1C CRP: c-reactive protein

## Ethical Statement

This study was exempt from review by the Institutional Review Board as we analyzed the NHANES cohort, which is considered secondary, publicly available, and de-identified data.

## Acknowledgements

N/A

## Funding, grant/award info

AG is supported by American Heart Association-Strategically Focused Research Network Grant in Disparities in Cardio-Oncology (#847740, #863620) and Department of Defense Prostate Cancer Research Program’s Physician Research Award (#HT94252310158).

## Supplemental

### Supplemental Methods

#### National Health and Nutrition Examination Survey

NHANES participants are interviewed at home as well as attend an examination site, where they undergo a variety of physical examinations, questionnaires, and blood sample testing. Questionnaires capture information about the participant’s demographics, diet, socioeconomic status, as well as health conditions and medications. With the physical exam and blood sample testing, NHANES attempts to capture clinical measurements such as blood pressure, body mass index, serum glucose, creatinine, and other biomarkers (Shay et al., 2013). For this analysis, data from NHANES years 1999-2010 was linked to the 2019 National Death Index (NDI), published by the National Center for Health Statistics (NCHS). This linkage was done by NCHS researchers matching adult NHANES participants to their mortality status using identifiers such as social security number, name, and date of birth. Mortality status was found using death certificates, information from the Social Security Administration, and Centers for Medicare and Medicaid Services. The causes of death were converted to the International Statistical Classification of Diseases, Injuries, and Causes of Death (ICD-10) codes. The NHANES-NDI linkage file that was used also stratified deaths by the nine leading causes of death, including , cerebrovascular disease, and heart disease.

**Supplemental Table 1:**
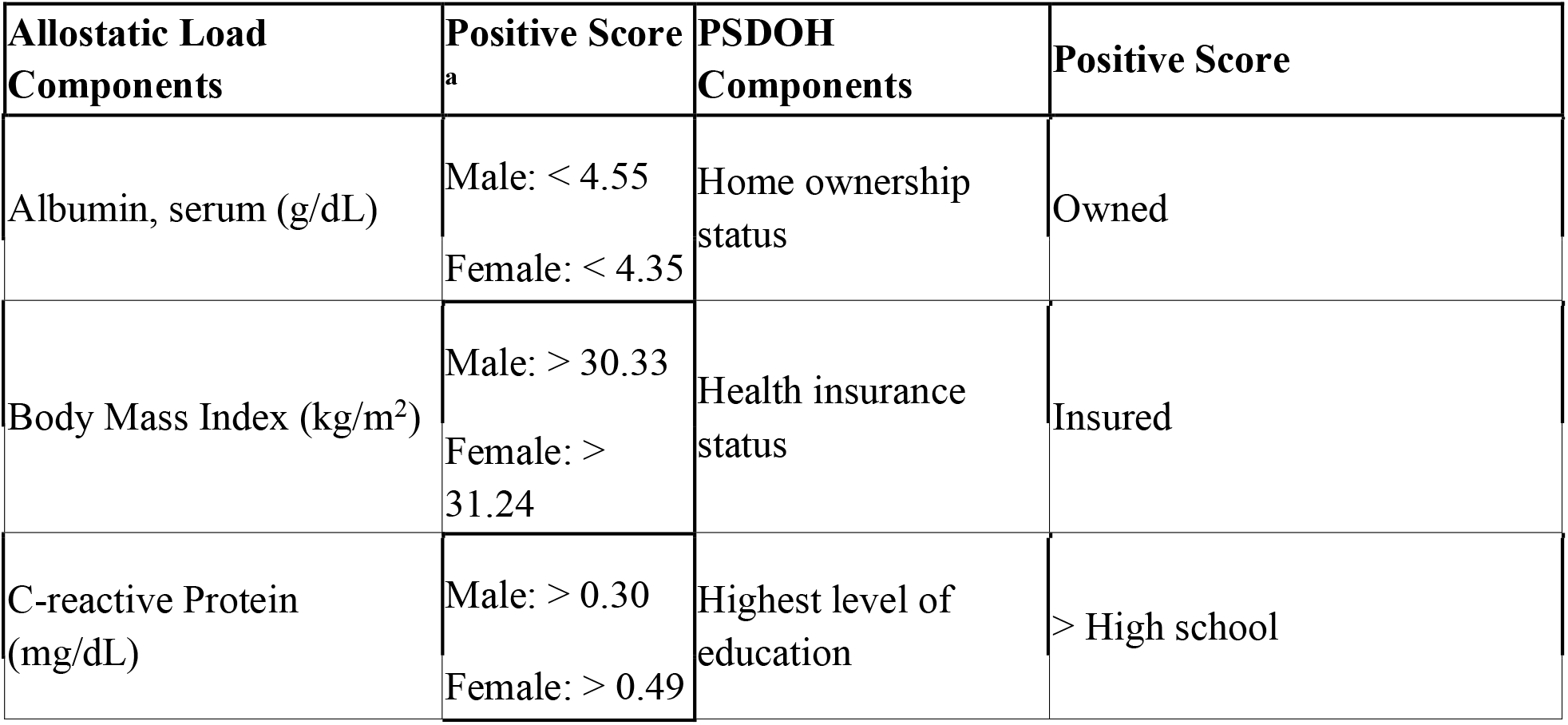
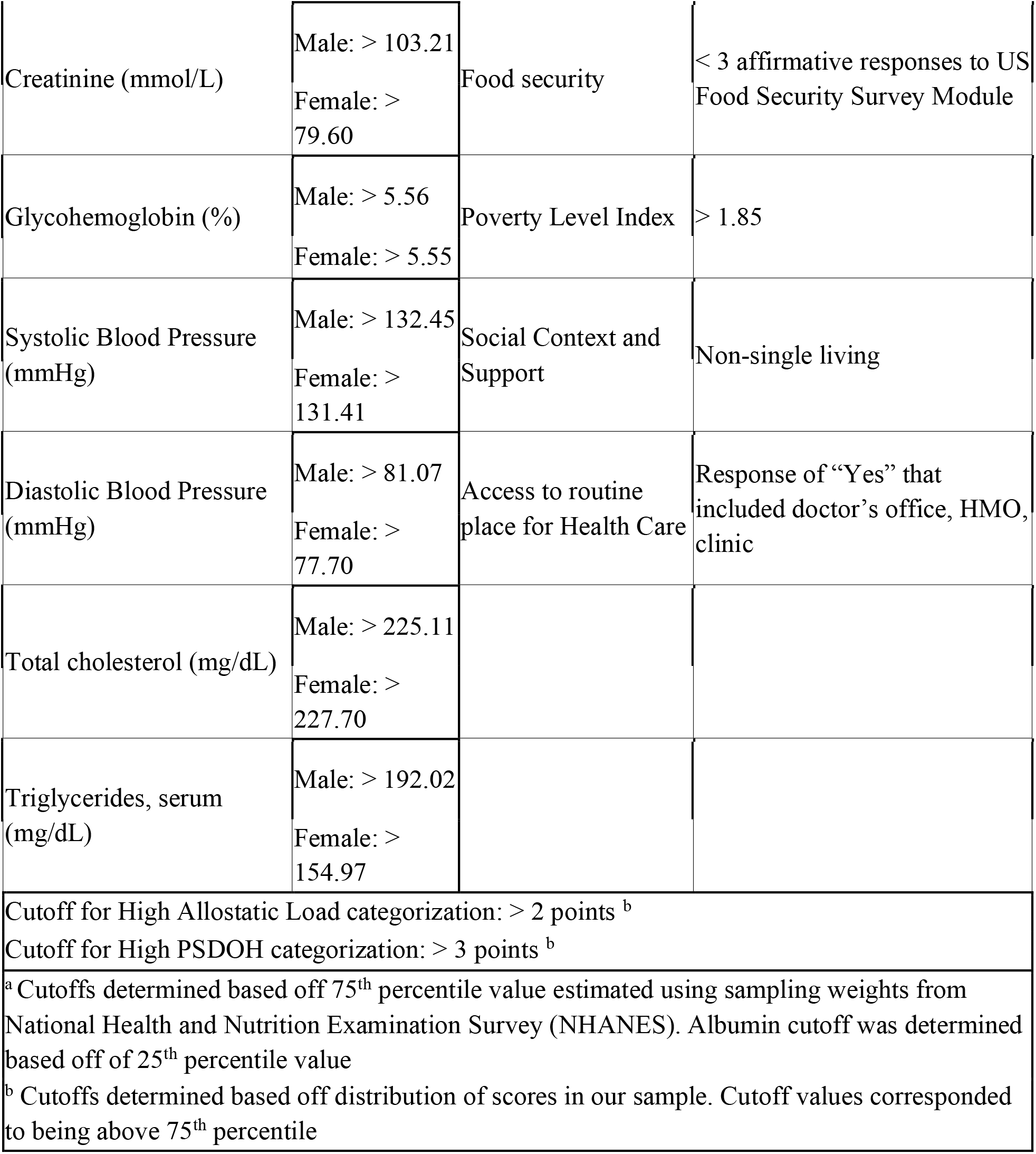
Component breakdown for calculation of Allostatic Load and Positive Social Determinants of Health scores.

**Supplemental Table 2.** Socio-demographic and medical descriptive characteristics among 20,836 National Health Examination Survey (NHANES) participants with no history of cancer stratified by AL status, years 1999-2010 with follow up through 2019.

| **No Cancer Incidence (N = 20836)** |  |  |  |
| --- | --- | --- | --- |
|  | **High Allostatic Load**<br>**(N = 9104)** | **Low Allostatic**<br>**Load (N = 11732)** | **P-Value** |
|  | Presented as N(%) or<br>Mean (95% CL) |  |  |
| **Allostatic Load Total Score** | 4.1 (0.012) | 1.0 (0.008) | p < 0.01 |
| **Sex** |  |  |  |
| Female | 4293 (47.2) | 5832 (49.7) | p < 0.01 |
| Male | 4811 (52.8) | 5900 (50.3) | p < 0.01 |
| **Mean Age in Years** | 54.7 (0.173) | 39.2 (0.161) | 0.0247 |
| **Race/Ethnicity** |  |  |  |
| Non-Hispanic White | 4226 (46.4) | 5724 (48.8) | 0.0194 |
| Non-Hispanic Black | 2105 (23.1) | 2029 (17.3) | p < 0.01 |
| Mexican-American/Other Hispanic | 2459 (27.0) | 3438 (29.3) | p < 0.01 |
| Other/ Mixed Race | 314 (3.45) | 541 (4.61) | 0.0213 |
| **SDOH score** |  |  |  |
| 0 | 43 (0.472) | 92 (0.784) |  |
| 1 | 210 (2.31) | 511 (4.36) |  |
| 2 | 669 (7.35) | 1181 (10.1) |  |
| 3 | 1255 (13.8) | 1692 (14.4) |  |
| 4 | 1549 (17.0) | 1847 (15.7) |  |
| 5 | 1838 (20.2) | 2005 (17.1) |  |
| 6 | 1970 (21.6) | 2325 (19.8) |  |
| 7 | 1570 (17.2) | 2079 (17.7) |  |
| **PSDOH level** |  |  |  |
| Low PSDOH | 2177 (23.5) | 3476 (30.3) | p < 0.01 |
| High PSDOH | 6927 (76.5) | 8256 (69.7) | p < 0.01 |
| **Time Period/Year** |  |  |  |
| 1999-2000 | 984 (10.8) | 1567 (13.4) | p < 0.01 |
| 2001-2002 | 1325 (14.6) | 2004 (17.1) | p < 0.01 |
| 2003-2004 | 1306 (14.3) | 1838 (15.7) | 0.0723 |
| 2005-2006 | 1465 (16.1) | 1963 (16.7) | 0.258 |
| 2007-2008 | 2030 (22.3) | 2029 (17.3) | p < 0.01 |
| 2009-2010 | 1994 (21.9) | 2331 (19.9) | 0.0266 |
| **Education** |  |  |  |
| Less than high school | 3125 (34.3) | 3272 (27.9) | p < 0.01 |
| High school or equivalent | 2294 (25.2) | 3179 (27.1) | p < 0.01 |
| Some college or AA degree | 2321 (25.5) | 2875 (24.5) | 0.2682 |
| College Graduate or more | 1356 (14.9) | 2397 (20.4) | p < 0.01 |
| **Income Relative to Federal Poverty Line** |  |  |  |
| 1st Quartile (0-1.11) | 2177 (23.9) | 2917 (24.9) | 0.9005 |
| 2nd Quartile (1.11-2.08) | 2402 (26.4) | 2724 (23.2) | p < 0.01 |
| 3rd Quartile (2.08-3.77) | 2064 (22.7) | 2657 (22.6) | 0.7687 |
| 4th Quartile (3.77-11.89) | 2461 (27.0) | 3434 (29.3) | p < 0.01 |
| **Current Smoker Status** | 1945 (21.4) | 2520 (21.5) | 0.064 |
| **Ever Congestive Heart Failure** | 377 (4.14) | 106 (0.90) | p < 0.01 |
| **Ever Heart Attack** | 530 (5.82) | 205 (1.75) | p < 0.01 |

**Supplemental Table 3.**
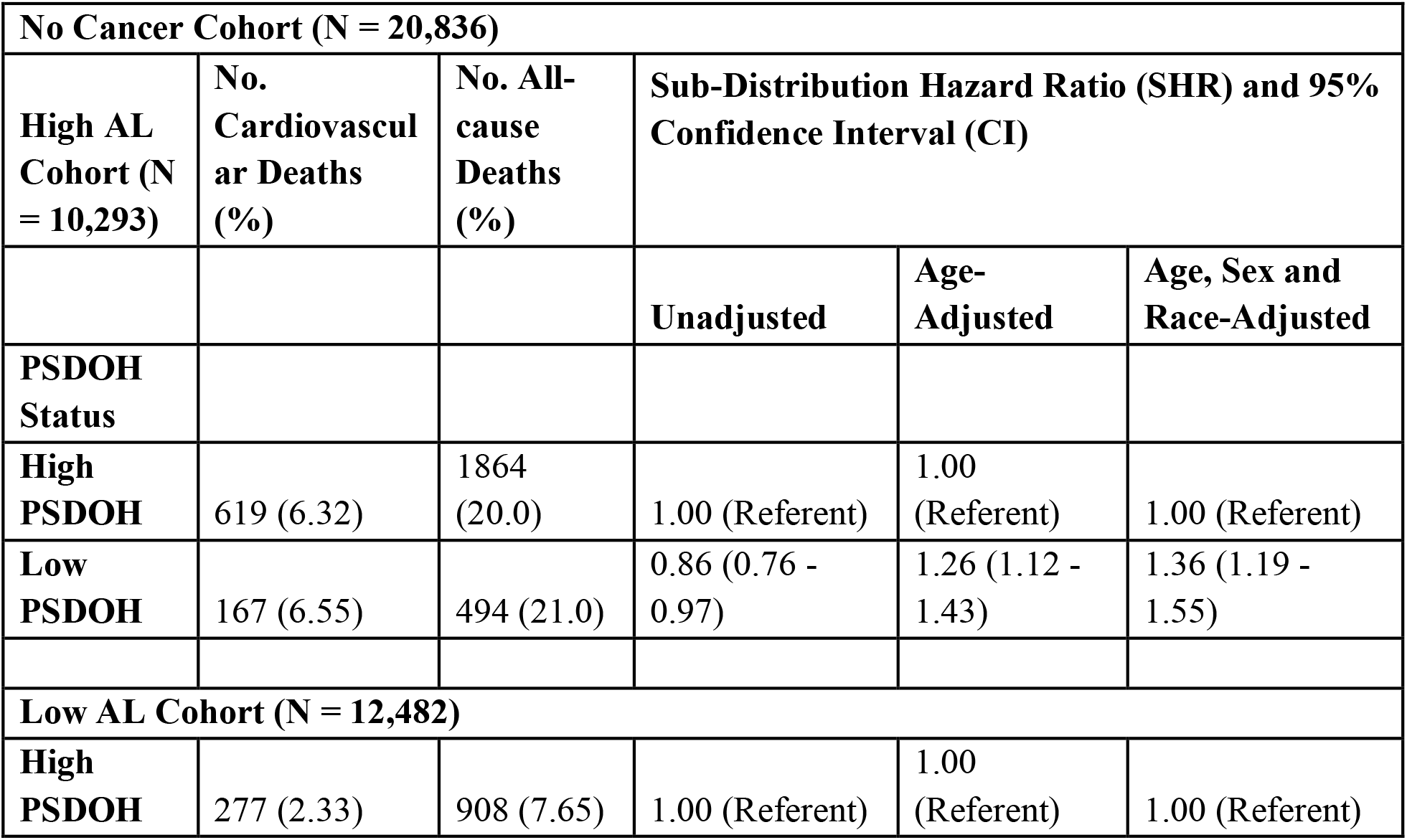
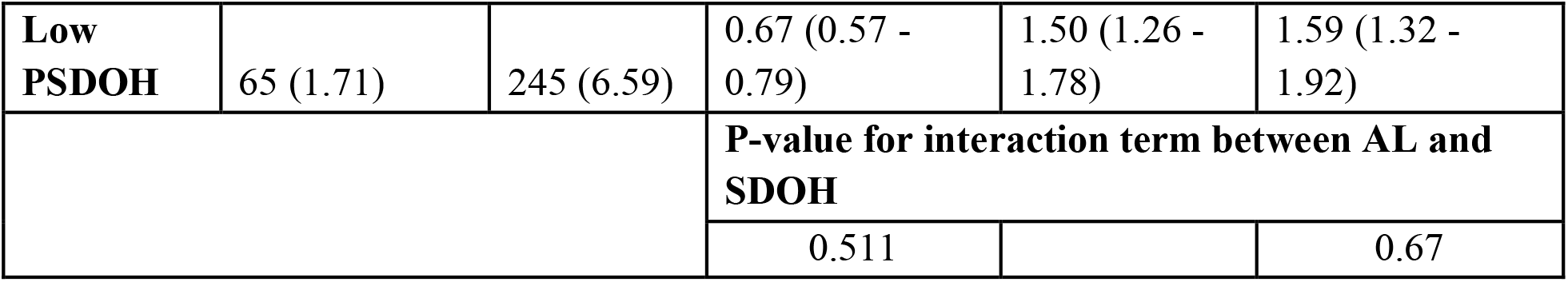
Fine & Gray method for proportional hazard models presented as Sub-Distribution Hazard ratios (SHR) and 95% confidence intervals (CI) for the association between PSDOH and CVD death among participants with no history of cancer stratified by AL status accounting for competing risks of all-cause mortality, among 20,836 NHANES participants with 1,128 CVD, and 3,511 competing deaths.

